# Data-independent acquisition phosphoproteomics of urinary extracellular vesicles enables renal cell carcinoma grade differentiation

**DOI:** 10.1101/2022.08.15.22278799

**Authors:** Marco Hadisurya, Zheng-Chi Lee, Zhuojun Luo, Guiyuan Zhang, Yajie Ding, Hao Zhang, Anton B. Iliuk, Roberto Pili, Ronald S. Boris, W. Andy Tao

## Abstract

Translating the research capability and knowledge in cancer signaling into clinical settings has been slow and ineffective. Recently, extracellular vesicles (EVs) have emerged as a promising source for developing disease phosphoprotein markers to monitor disease status. This study focuses on the development of a robust data-independent acquisition (DIA) using mass spectrometry to profile urinary EV phosphoproteomics for renal cell cancer (RCC) grades differentiation. We examined gas-phase fractionated (GPF) library, direct DIA, forbidden zones, and several different windowing schemes. After the development of a DIA mass spectrometry method for EV phosphoproteomics, we applied the strategy to identify and quantify urinary EV phosphoproteomes from 57 individuals representing low-grade clear cell RCC, high-grade clear cell RCC, chronic kidney disease (CKD), and healthy control (HC) individuals. Urinary EVs were efficiently isolated by functional magnetic beads and EV phosphopeptides were subsequently enriched by PolyMAC. We quantified 4,057 unique phosphosites and observed that multiple prominent cancer-related pathways, such as ErbB signaling, renal cell carcinoma, and regulation of actin cytoskeleton, were only upregulated in high-grade clear cell RCC, while those correlated with a higher survival rate were elevated in low-grade clear cell RCC only. These results show that EV phosphoproteome analysis utilizing our optimized procedure of EV isolation, phosphopeptide enrichment, and DIA method provides a powerful tool for future clinical applications.

## INTRODUCTION

Renal cell carcinoma (RCC) is currently the 8^th^ leading cause of cancer death in the United States, affects nearly 300,000 individuals worldwide each year, and is responsible for over 100,000 deaths annually (1, 2). RCC originates from the renal cortex or the renal epithelial cells and accounts for over 90 percent of all kidney cancers (3, 4). In the past decades, the incidence of RCC has been increasing steadily, and a diverse set of RCC subtypes has been recognized. The primary histologic subtypes are clear cell (70-80%), papillary (15%), chromophobe (5%) and unclassified RCC (5, 6). Distinct cytogenetic and immunohistochemical profiles characterize each subtype and prognoses as reflected by staging severity, with the lower stage being associated with longer survival rates (7). Clear cell RCC is the most common among these subtypes and accounts the majority of RCC-related deaths. Due to the lack of symptoms until locally advanced or metastatic, renal cell cancer is typically detected incidentally when localized without warning. Currently, the detection and classification of renal masses rely on radiologic examinations, including ultrasound (US), computed tomography (CT), magnetic resonance imaging (MRI), etc. (8). In recent years, the frequent use of imaging for unrelated clinical symptoms of other diseases has led to a higher number of incidental diagnoses of RCC (9). Once identified the majority of renal masses are operated on without knowledge of subtype or grade. There are a number of explanations for this approach including a high number of historical non diagnostic results, the risk of tumor seeding, and the risk of complications including primarily bleeding and pain as well as limited access to quality interventional radiology (10–14). Although recently renal mass biopsy has improved and utilization has increased, having an alternative office-based test that could predict tumor type, aggressiveness, and the need for surgical intervention while obviating the need for biopsy has been the elusive “holy grail” for the practicing urologist (8, 15). Considering that most of the identified tumors are low-grade, the alternative approach could allow the urologists to make decisions on a case-by-case basis depending on the grades of cancer, which might require active surveillance and different therapeutic strategies instead of surgical procedures.

Considering the limitations of current approaches, it is necessary to develop a novel diagnostic technique for early intervention of RCC. Therefore, early diagnosis and identification of RCC subtypes and tumor grades are essential to provide proper and effective treatment to increase the survival rate of patients. Recent studies suggest that extracellular vesicles (EVs) found in biofluids, such as urine, plasma, and saliva, can be a promising source for disease diagnosis (16–18). As shown in **Figure 1A**, EVs (e.g., exosomes and microvesicles) are membrane-covered particles containing bioactive molecules such as RNA, DNA, proteins, and lipids secreted by all types of cells that are crucial for cell-to-cell communications (19, 20). Exosomes are nanoscale vesicles ranging from 30–120 nm with spherical or cup-like morphology, whereas microvesicles are irregular in shape and tend to be larger with a wide range of sizes up to approximately 1,500 nm (21, 22). EVs secreted by cancer cells can promote cell growth and survival, shape the tumor microenvironment, and increase metastatic activities (23). Furthermore, EVs produced by cancer cells function as key mediators of cancer cell signaling and communication, causing adjacent or distant healthy cells to respond with phenotypic changes, which promote multiple aspects of tumor progression (23, 24). In addition, EVs are stably present in different body fluids, such as plasma and urine, which offers a useful and promising resource for non-invasive cancer biomarker discovery. In other words, EVs reflect the current disease state of cells by carrying bioactive components that can potentially be used as early biomarkers of RCC.

**Figure 1.**
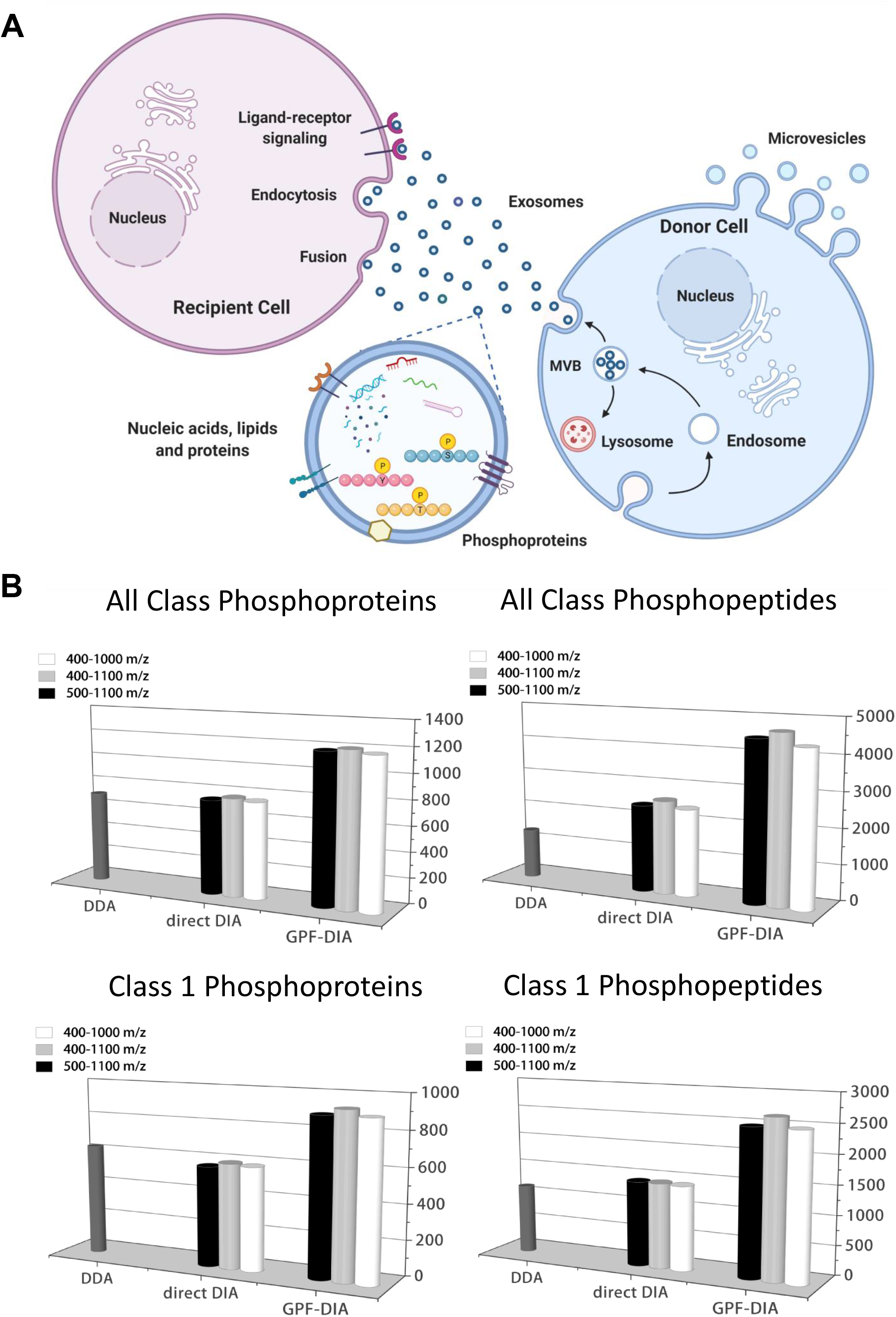
DIA method optimization. A) EVs, secreted from the parent cells, contain proteins, RNA, DNA, and lipids. Here, phosphoproteins directly reflect the cellular physiological status and signaling pathways during cancer progression. B) Top: The number of all class phosphoproteins (left) and phosphopeptides (right) in DDA, direct DIA, and GPF DIA (with three different m/z ranges). Bottom: The number of class 1 phosphoproteins (left) and phosphopeptides (right) in DDA, direct DIA, and GPF DIA (with three different m/z ranges).

Phosphoproteins in EVs offer valuable surrogates for monitoring disease states, as phosphorylation is a key regulator of proteins involved in different cellular functions such as cell growth, differentiation, and apoptosis (25). Alterations in phosphorylation pathways are often associated with devastating diseases such as cancer (25). Some well-known signaling pathways, such as MAPK, EGFR/HER, CDK, and Cadherin-catenin complex, are major cell cycle players and deregulation of their phosphorylation-dephosphorylation activity has been shown to lead to the formation of various types of cancers. Previous studies from our group have identified numerous EV phosphoproteins as potential disease markers in urine and plasma for breast cancer, chronic kidney disease, kidney cancer, and Parkinson’s disease patients (26–28).

A number of DIA methods have been introduced with increased coverage, sensitivity, and reproducibility compared to the more commonly used data-dependent acquisition (DDA) method and are powerful tools for disease biomarker discovery (29). This study aimed to present an EV phosphoproteomics approach based on optimized data-independent acquisition (DIA), by comparing different DIA strategies, including gas-phase fractionated (GPF) DIA library, direct DIA, forbidden zones, and several windowing schemes, to profile phosphoprotein landscape in urinary EVs from RCC patients and controls. Searle et al demonstrated that GPF DIA library has the advantage of ensuring that the library is experiment-specific, always up-to-date, and accounts for variation across different instrument platforms without the need of doing offline fractionation for library generation (30). Moreover, the narrow-window used by GPF-DIA provides parallel reaction monitoring (PRM) quality data for every peptide inside the scanned range. On the hand hand, direct DIA offers the benefit of performing DIA analysis without the need to build a library first. The windowing schemes were designed to be staggered, allowing peptide signal collection from multiple regions of the precursor space in the same MS2 and using the MS2 nearby in different precursor space regions to demultiplex signals specific to each region computationally. The demultiplexing confirms the presence of real peptide signals and eliminates potential noises. In addition, using an inclusion list with windows edged by forbidden zones, designed to lessen the quadrupole transmission edge effects, maximizes the precursor ion transmission in the window range. As a result, there was a slight improvement in phosphopeptide identification. Although our techniques were inspired by existing works, we made our own efforts to optimize the DIA method designed for urinary EV phosphoproteomics.

After we optimized the DIA phosphoproteomics method for urinary EV samples, we carried out DIA phosphoproteomics in urinary EVs derived from 57 individuals with low-grade clear cell RCC, high-grade clear cell RCC, chronic kidney disease (CKD), and healthy control (HC) to differentiate clear cell low-grade from CKD, controls, and clear cell high-grade RCC. We discovered that the GPF library and direct DIA combination provide the most comprehensive information for our clinical sample quantification. Some prominent cancer pathways, such as ErbB signaling, proteoglycans in cancer, renal cell carcinoma, and microRNAs in cancer pathways, were found to be upregulated only in the high-grade clear cell RCC urinary EV samples. Furthermore, some phosphosites were uniquely upregulated in either clear cell low-grade or high-grade RCC. High expression of SLC47A1, VPS37B, PDCD6IP, ARHGEF10L, PLEKHG3, and SOWAHC phosphosites exclusively found in the clear cell low-grade are prognostic, favorable in renal cancer, and are correlated with a higher survival rate based on the Human Protein Atlas (31). Meanwhile, several phosphosites known to be involved in RCC, such as PAK1 (S144, S174, T185), PAK2 S2, and BRAF (S365, S729), were upregulated in clear cell high-grade. Moreover, the renal cell carcinoma pathway was only enhanced in clear cell high grade RCC. These findings offer opportunities for further exploration to develop novel EV phosphoprotein-based biomarkers and open the door for an effective early-stage clinical diagnosis for clear cell RCC.

## EXPERIMENTAL PROCEDURES

### Sample Collection

Samples were collected at the Indiana University Simon Cancer Center and Methodist Research Institute in Indianapolis. The urine samples were collected on the same day right before the renal mass removal surgery. The collected urine samples were aliquoted into two or more cryotubes (5 mL) and stored in a -80ºC freezer. After the surgery, the obtained tumor tissue samples were used for immunohistochemistry (IHC) analysis to diagnose the cancer subtypes and the grade based on the WHO/ISUP grading system for RCC (32). We have collected diverse RCC subtypes; however, we only utilized the low-grade and high-grade clear cell RCC for this study. Some samples were collected and used for the method optimization. We also collected urine samples from non-cancer healthy individuals (HC) and chronic kidney disease (CKD) patients. Here, we were also interested in investigating whether CKD samples could serve as a better control group than HC group when they were utilized as a control group for clear cell low-grade and clear cell high-grade RCC differentiation. In total, we processed 60 samples (15 HC, 15 CKD, 15 clear cell low-grade, 15 clear cell high-grade) chosen randomly from a larger sample cohort. All samples were collected under IRB approved protocol no. 1011004282, Development of a Biorepository for IU Health Enterprise Clinical Research Operations. The study design and conduct complied with all relevant regulations regarding the use of human study participants and was conducted in accordance with the criteria set by the Declaration of Helsinki. All 60 samples were processed separately by implementing the statistical principles in experimental designs, including replication, randomization, and blocking when applicable (33).

### EV Isolation by EVtrap

The EVtrap beads were provided by Tymora Analytical Operations and were used as described previously (34). The 60 urine samples (approximately 10-15mL each) and the sample pool (60 samples combined for a total volume of 180 mL) were centrifuged at 2,500 x g for 10 min and frozen at -80°C. After thawing, the sample pool was split into 6 × 50 mL tubes (30 mL in each tube) to facilitate the EVtrap incubation. Magnetic EVtrap beads were added to each urine sample at the ratio of 20 µL beads per 1 mL of urine. The samples, including the sample pool, were then incubated for 1 hr by end-over-end rotation, allowing for ample movement of the beads. Afterward, the solution was removed with the aid of a magnetic separator. After three washes, the beads were incubated two times for 10 min by shaking with fresh 100 mM triethylamine to elute the EVs. After that, the eluted sample containing the EVs was collected, combined, and dried in a vacuum centrifuge.

### LC-MS Sample Preparation

The extracellular vesicles were solubilized with the help of a Phase Transfer Surfactant (PTS) buffer. The PTS buffer included 12 mM sodium deoxycholate (SDC), 12 mM sodium lauroyl sarcosinate (SLS), and 50 mM Tris-Cl, 10 mM tris-(2-carboxyethyl)phosphine (TCEP) to assist reduction, 40 mM chloroacetamide (CAA) to help alkylation, and phosphatase inhibitor to protect the phosphoproteins and phosphopeptides from phosphatases. 100 µL of PTS was added to each sample. The samples were boiled for 10 min at 95°C in the dark and were diluted five-fold using 50 mM TEAB for digestion. Then, a BCA assay was performed to identify the amounts of proteins per sample. Lys-C was added at the fixed 1:50 w/w ratio, and samples were digested at 37°C for 3 hours in a shaker at 1,100 rpm. Afterward, trypsin was added at the fixed ratio of 1 μg per 50 μg protein per sample, and samples were digested overnight at 37°C.

Each sample was acidified with 50 µL of 10% trifluoroacetic acid (TFA) solution to adjust its final concentration to 1% TFA. 650 µL of ethyl acetate was added to the sample and vortexed for 2 min to precipitate the detergents completely. The samples were then centrifuged at 20,000 x g for 3 min to differentiate the aqueous and organic phases. The upper layer (organic phase) was removed, and the samples were dried for about 1.5 hrs until less than 150 µL remained. Another 1 mL of ethyl acetate was added to the samples, and the previous steps of vortexing and centrifugation were repeated. This time, the samples were dried completely.

TopTip C-18 (10-200 µL) (Glygen, Part number: TT2C18.96) tips were used to desalt the samples according to the manufacturer’s instructions. Protein concentration was checked using a Pierce Colorimetry peptide concentration assay kit, normalized for the same amount of peptides for each sample, and dried completely. The sample pool utilized 50 mg Sep-Pak columns (Waters) for desalting according to the manufacturer’s instructions. After the concentration examinations using the Pierce Colorimetry peptide concentration assay, the appropriate peptide concentration for the GPF library (seven injections) was placed into several tubes (for PolyMAC preparation) and dried completely.

Each sample, including the pooled samples, was subjected to phosphopeptide enrichment using the PolyMAC Phosphopeptide Enrichment kit (Tymora Analytical) according to the manufacturer’s instructions (35). The eluted phosphopeptides were dried completely in a vacuum centrifuge.

### LC/MS-MS Analysis

The phosphoproteomic samples were spiked with an 11-peptide indexed Retention Time internal-standard mixture (Biognosys) to normalize the LC-MS signal between the samples. All samples were captured on a 2-cm Acclaim PepMap trap column and separated on a heated 50-cm Acclaim PepMap column (Thermo Fisher Scientific) containing C18 resin. The mobile phase buffer consisted of 0.1% formic acid in HPLC grade water (buffer A) with an eluting buffer containing 0.1% formic acid in 80% (vol/vol) acetonitrile (buffer B) run with a linear 85-min gradient of 5–35% buffer B at a flow rate of 300 nL/min. The UHPLC was coupled online with a Q-Exactive HF-X mass spectrometer (Thermo Fisher Scientific). For DDA experiments, the mass spectrometer was run in the data-dependent mode, in which a full-scan MS (from m/z 375 to 1,500 with the resolution of 60,000) was followed by MS/MS of the 15 most intense ions (30,000 resolution; normalized collision energy - 28%; automatic gain control target (AGC) - 2E4, maximum injection time - 200 ms; 60sec exclusion). For DIA experiments, the mass spectrometer was run in the data-independent mode, in which a full-scan MS (polarity - positive; scan range 389.8 to 1,109.8 m/z with the resolution of 60,000; automatic gain control target (AGC) - 1E6, maximum injection time - 60 ms; spectrum data type - centroid) was followed by MS/MS with 8.0 m/z staggered-isolation windows schemes as described in Searle *et*.*al*. and Pino *et*.*al*. (polarity - positive; 15,000 resolution; normalized collision energy - 27%; AGC - 1E6, maximum injection time - 20 ms; loop count - 88; spectrum data type - centroid] (29, 36).

### Construction of GPF and direct DIA library

The GPF spectra library was generated using Spectronaut Pulsar search (Biognosys, v15, Switzerland) according to Searle *et*.*al*. and Pino *et*.*al*. (29, 36). In brief, we acquired the GPF spectra library (n = 7) from narrow mass ranges: 394.8–504.8, 494.8–604.8, 594.8–704.8, 694.8– 804.8, 794.8–904.8, 894.8–1004.8 m/z, and 994.8–1104.8 m/z at the MS1 level. For the MS2 level, 4 m/z DIA spectra were acquired for each MS1 range. The effective isolation window is only 2 m/z after deconvolution. All the DIA data were first searched against the human FASTA file from UniProt Swiss-Prot downloaded December 20, 2021 (78,120 total entries) with a default setting. Maximum of two missed cleavages were allowed for trypsin digested peptides with variable modification as acetylation on protein N terminus (+42.016 Da) and oxidation of methionine (+15.995 Da) residues, as well as phosphorylation (+79.966 Da) on Ser, Thr, and Tyr residues. Fixed modification of carbamidomethylation (+57.022 Da) of cysteine residues was included. The tolerances for both MS1 and MS2 levels were set to dynamic (determined by Spectronaut based on the extensive mass calibration) with a correction factor of 1 (no correction). A cutoff of 1% FDR was set at both PSMs and peptides, and a cutoff of 5% FDR at the protein level with an enabled phosphosite localization filter at 0.75. Fragment ions minimum m/z 300, maximum m/z 1800, the minimal relative intensity of 5%, and 15 most intense fragment ions per precursor were included, and those with less than three amino acid residues were not considered. Fragment ions with neutral losses were included. The empirical iRT database was used for the iRT reference strategy with deep learning-assisted iRT regression as a backup. Precursors with phosphorylation modification as well as non-modified were retained. The library search for the direct DIA was established using the same parameters as the GPF spectral library described above (37).

### DIA Method Optimization

The placing of the window edges at the forbidden zones for phosphorylation PTM was performed and compared to without the forbidden zones using a precursor range of 400-1000 m/z. Then, three different precursor ranges (400-1000, 500-1100, and 400-1100 m/z) were compared. Finally, we compared two different methods for DIA library generation: the gas-phase fractionated (GPF) and direct DIA methods.

### Experimental Design and Statistical Rationale

During DIA method optimization, we focused on interpreting results from DDA and different DIA acquisition strategies, rather than any biological interpretation. As such, a single urine phospho-sample was injected to the mass spec with DDA and different DIA acquisition strategies (one MS run for each acquisition strategy). The different acquisition strategies were compared in terms of their phosphopeptides identification. For the application of the optimized method, we processed 60 samples (15 HC, 15 CKD, 15 clear cell low-grade, and 15 clear cell high-grade biological replicates) by implementing the statistical principles in experimental designs, including replication, randomization, and blocking when applicable (33). Here, CKD was used as another control group. Before MS analysis, indexed Retention Time (iRT) Standard containing 11 artificial synthetic peptides was added to the phosphoproteomic samples. MS2 level quantitation was obtained. During the MS data analysis, three samples (one CKD, one clear cell low-grade, and one clear cell high-grade) were eliminated from further analysis, where one had deficient identifications and the other two did not meet normal distribution requirements. All abundances, which have normal distributions, were median-normalized for each sample, and p-value controlled Welch’s two-sample t-test was performed for each of the comparisons.

### DDA Data Analysis for Identification

The DDA raw file was searched directly against the human FASTA file from UniProt Swiss-Prot downloaded December 20, 2021 (78,120 total entries) with no redundant entries, using Sequest search engines in Proteome Discoverer 2.3 software (Thermo Fisher Scientific). The mass tolerance was set at 10 ppm for MS1 and 20 ppm for MS2. In the processing workflow, search criteria for the search engine were performed with full trypsin/P digestion, a maximum of two missed cleavages allowed on the peptides analyzed from the sequence database, carbamidomethylation on cysteines (+57.0214 Da) as a static modification, and oxidation (+15.9949 Da) on methionine residues, acetylation (+42.011 Da) at N terminus of proteins, and phosphorylation (+79.996 Da) on serine, threonine, or tyrosine residues as variable modifications. The false-discovery rates (FDR) of proteins and peptides were set at 0.01. All protein and peptide identifications were grouped, and any redundant entries were removed. Phosphosites localization tool was applied to filter class 1 localization (class 1 probability cutoff ≥ 0.75). The total identification numbers of unique phosphopeptides and unique master phosphoproteins were reported.

### DIA Data Analysis for both Identification and Quantification

The signal extraction and quantitation of the DIA data were performed in Spectronaut™ (Biognosys, v15), utilizing a standard setting with some modifications. Briefly, dynamic retention time prediction with local regression calibration was used. MS1 and MS2 interference corrections were enabled. The FDR at peptide precursor was set to 1%. Meanwhile, the protein level was analyzed with scrambled decoy generation and dynamic size at 0.1 fractions of library size. The quantitation was performed at the MS2 level by enabling local cross-run normalization. Qvalue percentile with 0.2 fraction and no imputing was used as data filtering. Phosphopeptide precursor abundances were measured by the sum of fragment ion peak areas, and peptide grouping was performed based on modified peptides. Phosphosites localization tool was applied to filter class 1 localization (class 1 probability cutoff ≥ 0.75 for libDIA, and ≥0.99 for direct DIA). For site-specific quantitation, the abundance of a phosphorylation site was calculated by summing up all of the abundances in precursors/peptides, which contain the site in the corrected precursor/peptide abundance file. Customized R scripts adapted from Kitata *et*.*al*. performed site-specific quantification.

### Statistical analysis and pathway annotation

Further statistical analysis was performed by Perseus software (1.6.15.0) for the 60 urine samples (38). Those values with raw intensities below 30 were converted to NaN for quality control. All the phosphosite abundances were log2 transformed. The abundances were grouped into four different groups: HC, CKD, clear cell low-grade, and clear cell high-grade. Then, three samples (one CKD, one clear cell low-grade, and one clear cell high-grade) were eliminated from further analysis because they did not satisfy the abundance normal distribution requirement. The other 57 samples were further split into five comparisons: CKD vs. HC, clear cell low-grade vs. HC, clear cell high-grade vs. HC, clear cell low-grade vs. CKD, and clear cell high-grade vs. CKD. The phosphosites with more than 70% quantified abundances in at least one category were kept for each comparison. Imputation for the missing abundances was performed by assigning small values from the normal distribution (1.8 SDs downshift and 0.3 SDs width). Then, all abundances were further median-normalized for each sample. P-value controlled Welch’s two-sample t-test was performed for each of the comparisons (with cutoff values of p-value = 0.05 and log base 2 difference = 0.5, which equals ∼ 1.414 fold-change). Protein-protein interaction network analysis and functional annotation were done by STRING database (version 11.0b) (39) and Ingenuity Pathway Analysis (IPA) (40). For this analysis, pathways from the biological processes, molecular functions, and KEGG (41) databases were considered.

## RESULTS

### Exploration of DIA measurement strategy

The reproducibility and sensitivity of data-dependent acquisition (DDA) mass spectrometry is highly dependent on sample complicity and instrument conditions, and therefore, it is often less suitable for biomarker discovery research. To overcome DDA limitations, DIA has been developed that sliced the peptide ion space into segments for MS2 measurement to counterbalance the complexity of biological samples. DIA has been shown to improve reproducibility, quantitative precision, and proteome coverage compared to label-free DDA. The overall goal of using DIA in this study was to measure as much phosphoproteome as possible while maintaining quantitative accuracy. The balance between these two goals is critical for successful DIA experiments. To accomplish this for our study, we focused on several objectives: (a) maximize the precursor ion transmissions, (b) maximize the total precursor range of targeted phosphopeptides, and (c) generate the most efficient DIA library.

The placing of forbidden zones for particular post-translational modifications (PTMs), particularly phosphorylation, has been discussed previously, but it was never tested with actual phospho-samples (29). For the first objective, we placed the window edges at the forbidden zones for phosphorylation PTM and discovered that the window edge placing slightly increased the phosphopeptide identification (**Supplementary Table 1**). As predicted, the slight increase was due to less pronounced quadrupole transmission edge effects. Furthermore, the most optimal precursor range for phosphopeptide precursors was suggested to be 500-1100 m/z. For the second objective, we compared different precursor ranges (400-1000, 500-1100, and 400-1100 m/z) and found that the 400-1100 m/z precursor range was the most ideal for phosphoprotein and phosphopeptide detection (**Fig. 1B**, see **Supplementary Table 2** for a table format).

Lastly, a library must be generated before analyzing the data to overcome the complexity of DIA data. Typically, libraries are built from fractionated DDA data at the cost of significant additional work, loss of sample, and instrument time. These obstacles, along with dependence on the quality of library, make DDA-based library generation less suitable for more dynamic phosphoproteome clinical samples. Recently, several alternative approaches have been developed to overcome those challenges by building DIA-only chromatogram libraries, such as the gas-phase fractionated (GPF) and direct DIA methods (29, 36, 37). To our knowledge, there have not been published reports comparing the two methods for phosphoproteome data prior to this study. Using GPF DIA and/or direct DIA methods can help eliminate the potential problems with the DDA-based spectral libraries that are not reasonable representations of the DIA MS/MS. Therefore, to accomplish the last objective, we compared the identification results when the GPF library and direct DIA were used for analyzing DIA data. Together, we find that by optimizing the total precursor ranges, setting the window edges at forbidden zones, and using the GPF library for a single shot mass spectrometry run of an EV sample, Spectronaut identified 88.8% (direct DIA) and 237% (GPF DIA) more of all-class phosphopeptides compared to DDA analyzed by Sequest alone (**Fig. 1B**, see **Supplementary Table 2** for a table format). For class 1 phosphopeptides, we identified 23.2% (direct DIA) and 126% (GPF-DIA) more phosphopeptides than with DDA. We also found that most all-class phosphoproteins were class 1 phosphoproteins.

### GPF-DIA and direct DIA spectral libraries conjointly provide more comprehensive identification and quantitation results

After finding the best DIA method for our mass spectrometry data acquisition, we applied this optimized procedure to urine samples from 60 patients using approximately 10-15mL urine from each (**Fig. 2**). We also combined all 60 samples together and analyzed that pooled sample separately to create the GPF library. These 60 samples represented four groups: healthy control (HC), chronic kidney disease (CKD), low-grade clear cell RCC, and high-grade clear cell RCC. Clear cell renal cell carcinoma is the most common RCC subtype, accounting for 75-80% of sporadic cases (42). The study’s overall goal was to investigate whether our optimized method could be used to distinguish between low-grade and high-grade clear cell disease. In addition, we aimed to see whether CKD could be used as a better control group when investigating renal cell carcinoma biomarkers.

**Figure 2.**
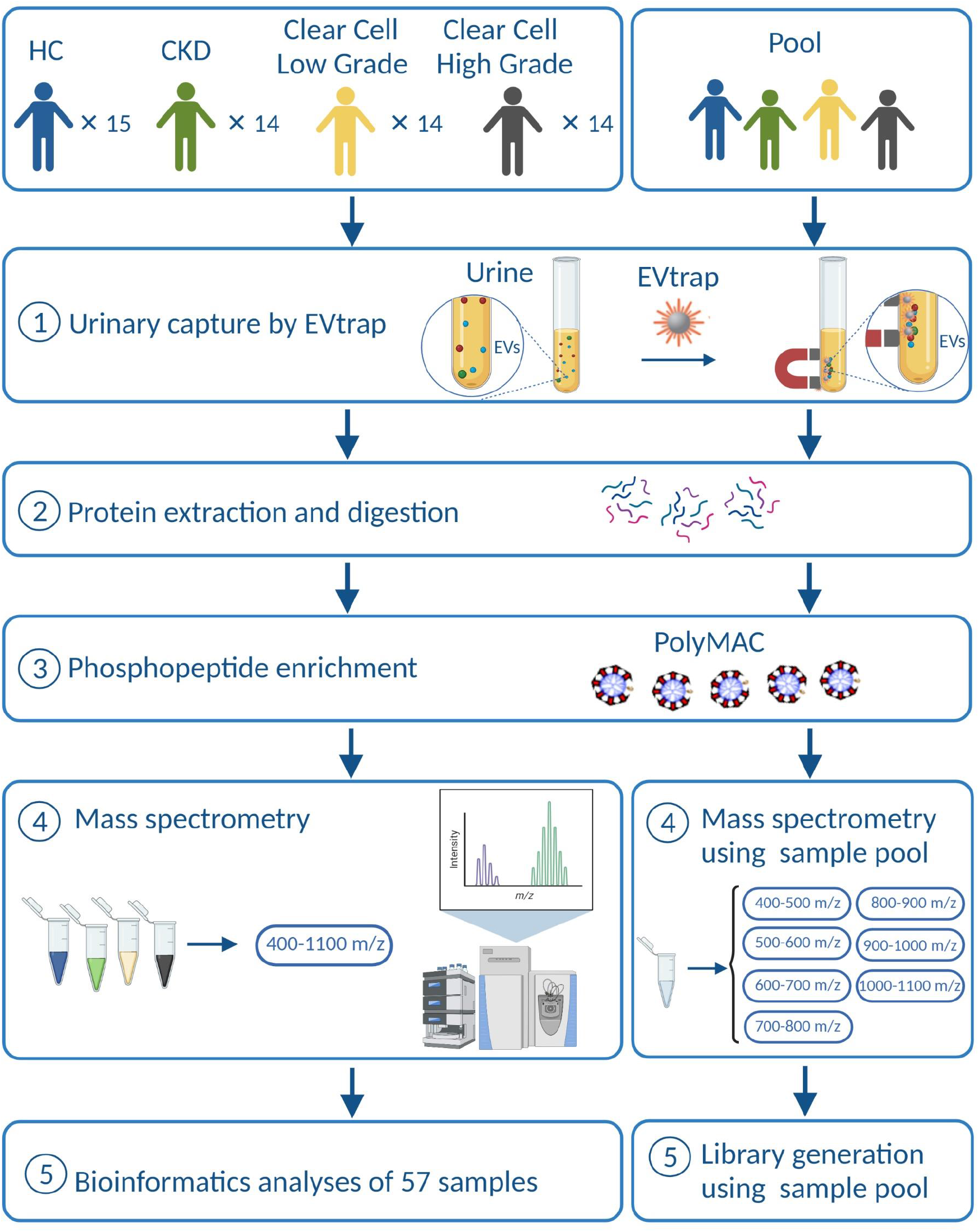
Schematic diagram of the EV biological features and the application of the optimized method. The workflow of the optimized DIA methods on 57 samples representing 4 different groups, HC (healthy control), CKD (Chronic Kidney Disease), Clear Cell Low-Grade RCC, and Clear Cell High-Grade RCC. The urine samples were processed using our in-house EVtrap for EV isolation and PolyMAC for phosphopeptides enrichment.

We collected the EVs from the urine samples using the synthesized magnetic beads (EVtrap) described previously (34). Following EV elution and drying, we lysed them with the optimized phase-transfer surfactant (PTS)-based procedure to extract and denature proteins. After the reduction/alkylation step, the proteins were digested with sequential Lys-C and trypsin additions, and the resulting peptides were desalted. We enriched phosphopeptides using the dendrimer-based PolyMAC method on each sample and analyzed them by LC-MS. Indexed Retention Time (iRT) Standard containing 11 artificial synthetic peptides was added to the phosphoproteomic samples to ensure calibration on difficult matrices and allow for detailed quality control readouts. The samples were analyzed by Thermo Fisher Q-Exactive HF-X MS coupled with the Ultimate 3000 UHPLC system. The same procedure was employed for the pooled sample for the GPF library generation.

High-quality and in-depth coverage spectral libraries are crucial for extensive DIA identification and quantitation. This is particularly important for EV samples because EV phosphoproteomics is only a small representation of total cell phosphoproteomics. In this case, we generated the GPF-DIA spectral library using the sample pool processed similarly to the individual samples. The sample pool was injected into the mass spectrometry seven times representing different MS1 window placements: 394.8–504.8, 494.8–604.8, 594.8–704.8, 694.8–804.8, 794.8–904.8, 894.8–1004.8 m/z, and 994.8–1104.8 m/z. For the MS2 level, staggered 4 m/z DIA spectra were acquired for each MS1 range giving the effective isolation window of only 2 m/z after deconvolution. The Pulsar search engine in the Spectronaut was used to generate the GPF library. We identified 9,559 all-class phosphopeptides and 2,441 all-class phosphoprotein groups (8,459 class 1 phosphopeptides and 2,409 class 1 phosphoprotein groups) (**Fig. 3A**). The kinase tree revealed 130 kinases identified in the library (**Fig. 3B**). Throughout this study, the data analyzed by the GPF library were denoted GPF-DIA. Since Spectronaut does not require a spectral library to analyze DIA data, we also performed a search using the directDIA function (37). Throughout this study, the data analyzed using the directDIA function were denoted as direct DIA.

**Figure 3.**
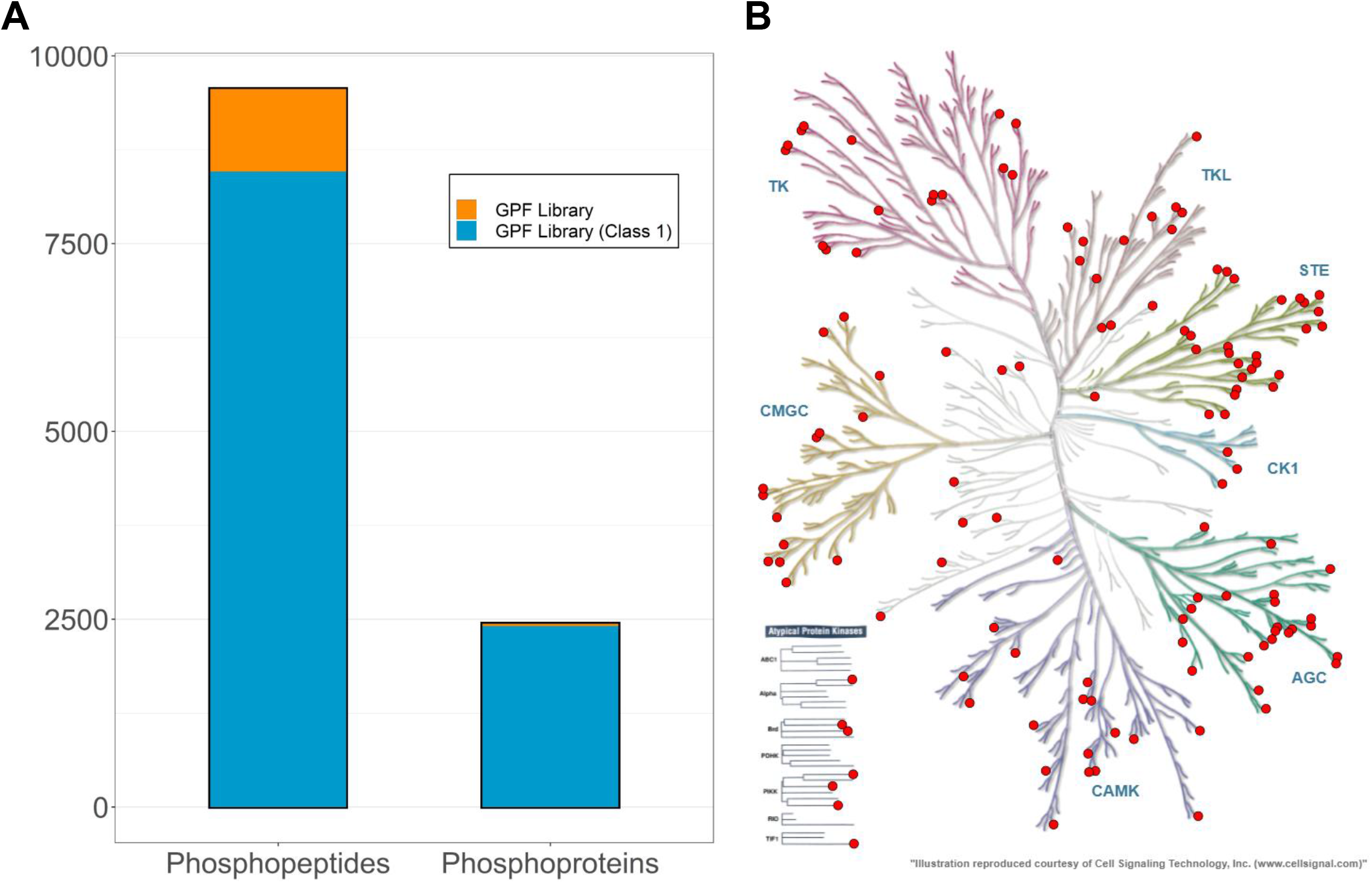
Composition of the GPF library for the EV phosphoproteome system. A) The number of phosphoproteins and phosphopeptides in the GPF library. B) Kinase tree revealed 130 kinases in the GPF library. KinMap database (http://www.kinhub.org/kinmap/) was used with input of protein accession number from GPF library. The kinase families listed include TK (tyrosine kinases), TKL (tyrosine kinase-like), CK1 (casein kinase 1), CAMK (calcium/calmodulin-dependent protein kinase), AGC (containing PKA, PKG, PKC families), CMGC (containing CDKs, MAPK, GSK, CLK families), and STE (serine/threonine kinases many involved in MAPK kinases cascade).

The quantified class 1 features were visualized for every individual, where we could observe consistent numbers of phosphoproteins across 57 samples (**Fig. 4A and 4B**). Using the GPF-DIA method, our urinary EV phosphoproteomic analyses quantified 1,417 unique phosphoproteins, 3,384 unique phosphopeptide groups, and 2,883 unique phosphosites (**Fig. 4C**). Meanwhile, direct DIA enabled the identification of 1,472 unique phosphoproteins, 3,585 unique phosphopeptide groups, and 2,988 unique phosphosites. Although we have shown that GPF DIA outperformed a single sample comparison against the direct DIA in Figure 1B, this was not the case anymore for multiple samples. The GPF DIA library was built from 7 different gas-phase fractionated MS runs from a single sample; therefore, the GPF DIA library generated a more comprehensive library than the direct DIA used to search for a single sample run. Regarding the phosphosite distribution in both GPF-DIA and direct DIA, we quantified about 78% pSer, 17% pThr, and 5% pTyr. The GPF-DIA and direct DIA provided 1,069 and 1,174 unique quantified phosphosites, respectively, enriching potential phosphoproteomes for RCC grade differentiation (**Fig. 4D**). The unique phosphosites found only in the GPF-DIA analysis arose from the simplified (fractionated) MS1 and MS2 data acquisitions during the generation of the GPF library. In contrast, the dilution of low-abundant phosphopeptides in the combined pool for GPF-DIA gave rise to those unique phosphosites only found in the direct DIA analysis. In other words, direct DIA overcame issues with GPF-DIA, where rare or low-abundant phosphorylation sites may get diluted out in the sample pool combined for library generation (37).

**Figure 4.**
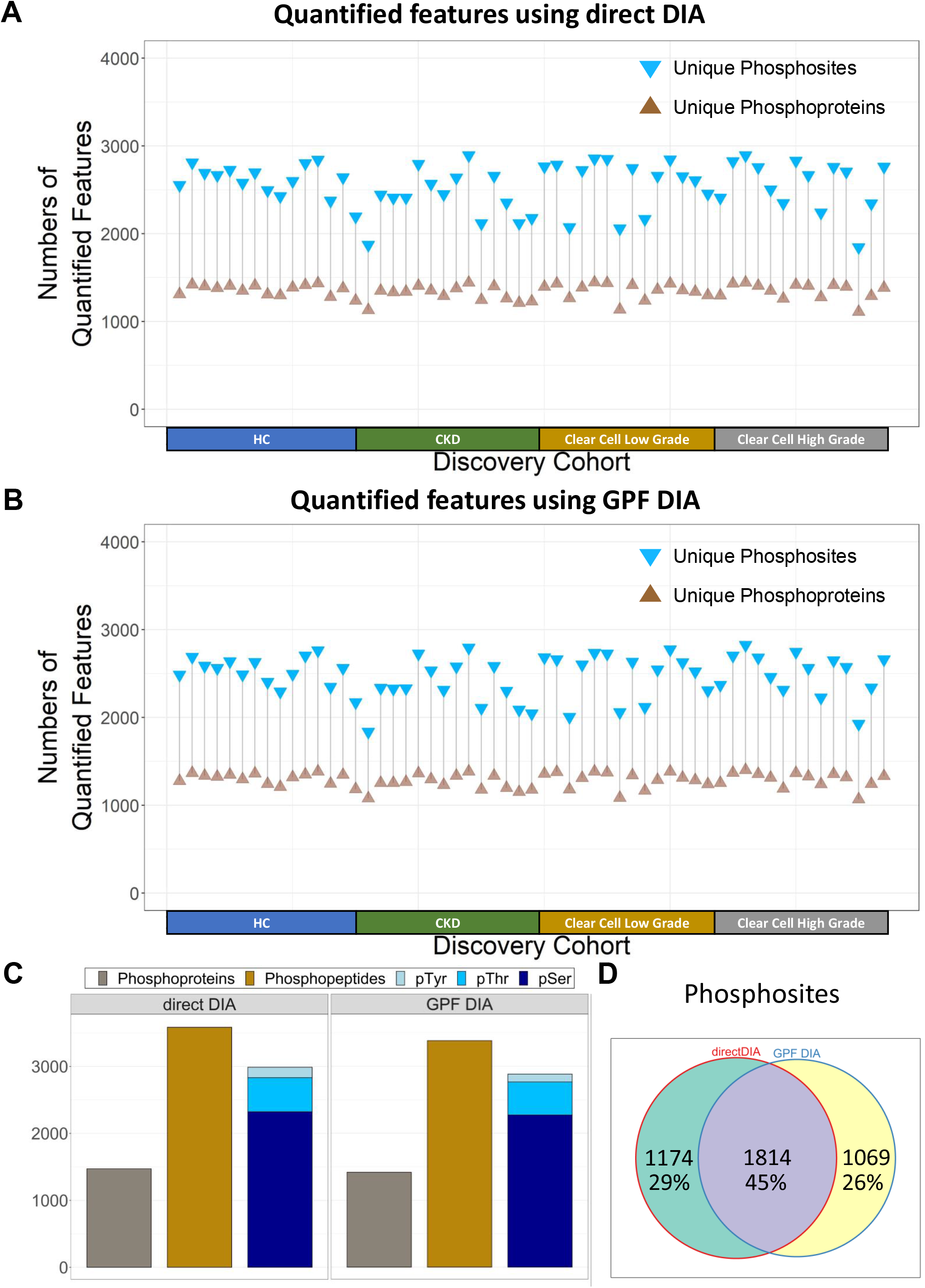
The summary of identification and quantification for all 57 patients. Cleveland Dot Plots for all quantified phosphoproteins and phosphosites searched using (A) direct DIA and (B) GPF DIA across all 57 patients. C) The number of quantified phosphoproteins and phosphopeptides searched using direct DIA and GPF library along with the distribution of quantified serine (pSer), threonine (pThr), and tyrosine (pTyr) phosphosites. D) The overlap of phosphosites quantified using direct DIA and GPF DIA (All quantified features are class 1).

Subsequently, we combined all unique phosphosites quantified in direct DIA and GPF-DIA because they would provide a more comprehensive picture of the cancer grades. First, we designated the phosphosites found in GPF-DIA with a unique label so that they can be differentiated from the phosphosites quantified in direct DIA. For our downstream analysis, we focused on those quantified 2,883 phosphosites using the GPF-DIA and the 1,174 phosphosites uniquely quantified in direct DIA. A total of 4,057 phosphosites were statistically analyzed in Perseus (1.6.15.0) (38).

### The renal cell carcinoma pathway is only upregulated in the high-grade clear cell RCC urine EVs

As described in the experimental procedure section, the quantified phosphosites must pass a rigorous statistical threshold and normalization criteria to be statistically useful. Out of 60, 3 samples failed to fulfill the requirements for statistical analysis, where one had deficient identifications and the other two did not meet normal distribution requirements. From a total of 57 samples, we then identified the upregulated phosphosites in CKD, low-grade and high-grade clear cell RCC groups against the healthy controls using volcano plots (with cutoff values of Welch’s two-sample t-test p-value = 0.05 and log base 2 difference = 0.5, which equals ∼ 1.414 fold-change, see **Supplementary Figure 1A**). We identified 514 upregulated phosphosites in CKD, 64 upregulated phosphosites in low-grade clear cell RCC, and 102 upregulated phosphosites in the high-grade clear cell RCC (**Supplementary Figure 1C**). Interestingly, we discovered more upregulated phosphosites in the CKD vs. healthy control sample than in the other two comparisons. It is possible that these upregulated phosphosites are associated with the CKD patients’ immune responses.

The most statistically significant phosphosites in the low-grade and high-grade clear cell RCC samples are visualized in the heatmap in **Figure 5A** (p-value < 0.05 when low-grade was compared to HC, CKD, or high-grade and when high-grade was compared to HC, CKD, or low-grade, calculated using the unpaired two-samples Wilcoxon test). PAK1, one of the class I p21-activated kinases, is a serine/threonine kinase involved in such cellular activities as neurogenesis, angiogenesis, cell migration, cytoskeletal dynamics, mitosis, apoptosis, and transformation (43). Highly phosphorylated PAK1 is associated with poor prognosis in RCC patients (44, 45). Therefore, it is unsurprising that we found several phosphorylation sites on PAK1 (S144, S174, T185) to be upregulated in a high-grade clear cell RCC (**Fig. 5A**). Moreover, S144 is a known autophosphorylation important to PAK1 function and is associated with increased PAK1 activity (46). The phosphorylated PAK2 at S2 was also significantly upregulated in this sample group. As another class I p21-activated kinase, PAK2 has been identified as a novel activator of MAPK Erk3 signaling (47). Although there are no reports about the specific VCAN phosphorylation at S2116, clear cell RCC was shown to have an elevated expression of VCAN in both cell lines and clinical specimens (48). This study also revealed that there is a strong correlation between VCAN expression and metastasis (P<0.001) and worse 5-year survival after radical nephrectomy (P=0.014). Interestingly, BRAF phosphorylation at S365 and S729 act as 14-3-3 protein binding sites and inhibit the activation of BRAF kinase activity (49). Furthermore, several phosphosites were found to be upregulated only in the low-grade clear cell RCC urinary EVs (**Fig. 5A**). High expressions of SLC47A1, VPS37B, PDCD6IP, ARHGEF10L, PLEKHG3, and SOWAHC phosphosites as depicted in **Figure 5A** are prognostic, favorable in renal cancer, and are correlated with a higher survival rate based on the pathology atlas of the human cancer transcriptome of the Human Protein Atlas (31). The renal cancer data in the pathology atlas of the human cancer transcriptome comprises 877 (651 alive and 226 dead) individuals from all four stages of renal cancer.

**Figure 5.**
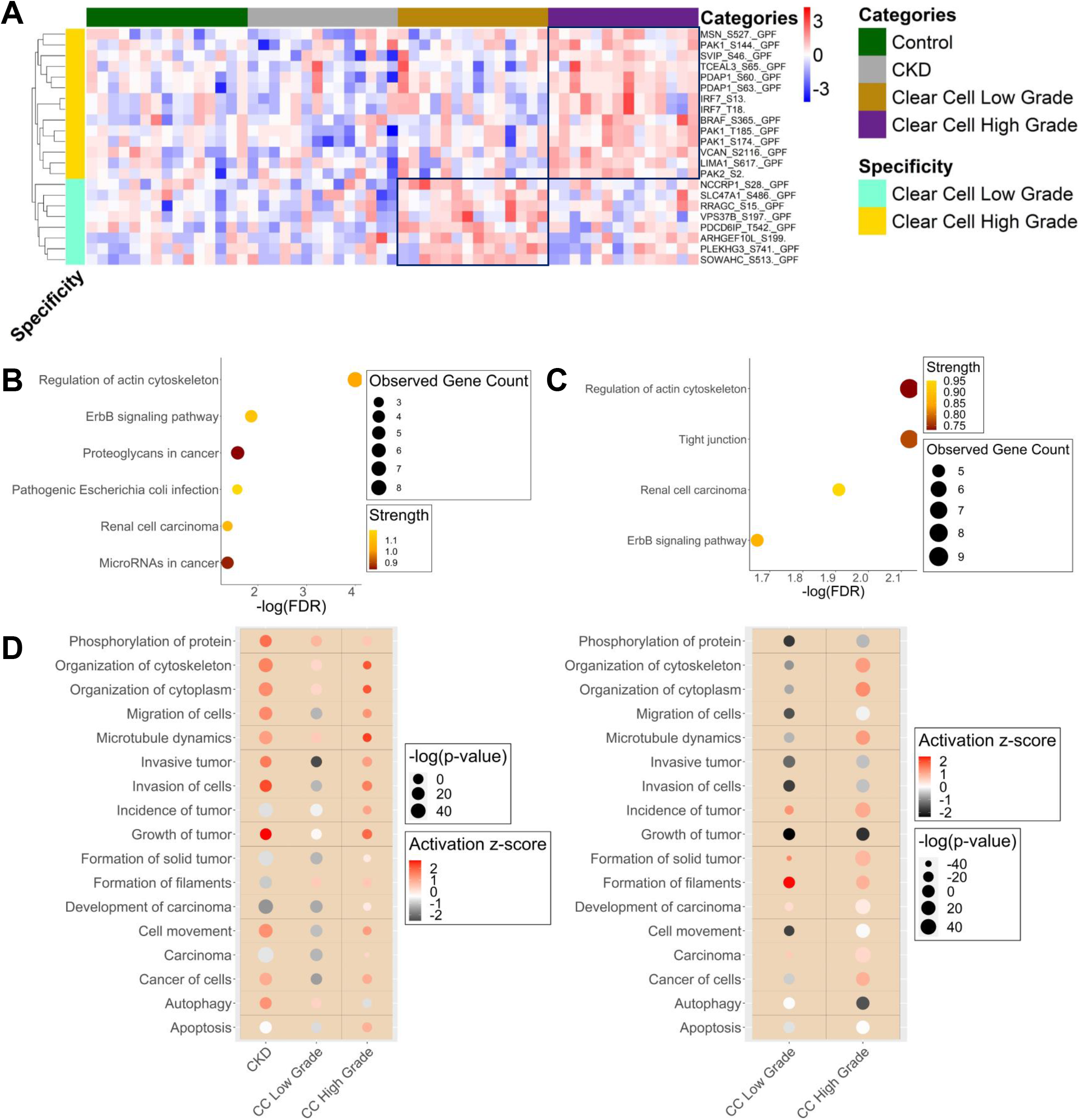
The RCC subtype differentiation. A) The heatmap of upregulated phosphosites in clear cell low-grade and high-grade RCC. Those phosphosites marked with GPF were quantified using the GPF library; meanwhile, those phosphosites without GPF were quantified using the direct DIA. B) The upregulated KEGG pathways in clear cell high grade RCC versus control. C) The upregulated KEGG pathways in clear cell high grade RCC versus CKD. D) Diseases and functions analysis on CKD, clear cell low-grade, and clear cell high-grade RCC versus the control group (left). Diseases and functions analysis on clear cell low-grade and clear cell high-grade RCC versus the CKD group (right)

Gene ontology analysis was performed on the upregulated phosphosites in String proteins to obtain data on biological processes, molecular function, and the Kyoto Encyclopedia of Genes and Genomes (KEGG). In CKD, mRNA-related pathways, exocytosis, leukocyte activation, immune system process, and cell secretion were upregulated, mediating the immune system and inflammatory responses (**Supplementary Figure 2A**). This likely explains our results showing increased upregulated phosphosite identification in CKD compared to RCC samples. The low-grade clear cell RCC GO analysis showed an upregulated junction assembly pathway (**Supplementary Figure 2B**). Meanwhile, the cytoskeleton organization-related pathways were found to be upregulated in the high-grade clear cell and are commonly dysregulated in cancer (**Supplementary Figure 2C**). Many high-grade clear cell molecular function pathways support the upregulation of cytoskeleton organization. For KEGG, the prominent cancer pathways, such as ErbB signaling, proteoglycans in cancer, renal cell carcinoma, and microRNAs in cancer pathways, were found to be upregulated only in the high-grade clear cell RCC urinary EV samples (**Fig. 5B**). The renal cell carcinoma pathway included some already known phosphoproteins involved in RCC, such as PAK1, PAK2, and BRAF (44, 50, 51). These three phosphoproteins are also involved in ErbB signaling and regulation of actin cytoskeleton pathways.

Next, we asked whether CKD was a better control than healthy individuals in our data analysis. We hypothesized that using the CKD as a control group could help eliminate the immune response from the more relevant cancer pathways analysis. We compared the upregulated phosphosites in clear cell RCC groups against the CKD cohort by using volcano plots (with cutoff values of Welch’s two-sample t-test p-value = 0.05 and log base 2 difference = 0.5, which equals ∼ 1.414 fold-change, see **Supplementary Figure 1B**). We identified 210 upregulated phosphosites in low-grade clear cell RCC and 227 upregulated phosphosites in the high-grade clear cell RCC samples (**Supplementary Figure 1D**). In this comparison, the endocytosis pathway was upregulated in the low-grade clear cell group (**Supplementary Figure 2D**). For KEGG, we observed similar pathways as previously shown when using the healthy controls as the control group. Most importantly, we observed a significant upregulation of the renal cell carcinoma pathway (**Fig. 5C**). The renal cell carcinoma pathway included the previously identified three phosphoproteins (PAK1, PAK2, and BRAF), as well as PAK4 and BAD phosphorylation. PAK4, together with NAMPT, has been targeted for a dual and specific inhibition to decrease RCC growth (52). Although the role of BAD phosphorylation is not well-studied in RCC, BAD acts as a pro-apoptotic protein in several other cancers (53).

We also compared our upregulated pathways with a large proteogenomic study based on 103 clear cell RCC tissue samples (54). In that study, the authors created thirty phosphoproteomics co-expression network-modules containing at least twenty genes in each and performed KEGG and Reactome (55) pathway enrichment analyses to identify biological pathways overrepresented in each network-module via Fisher’s exact test. Although these phosphoproteomics clusters lacked differentiation between clear cell low-grade and high-grade RCC, these clusters displayed a general picture about what pathways were upregulated in clear cell RCC. Not surprising that those pathways discussed above, such as endocytosis, ErbB signaling, proteoglycans, pathogenic *E. Coli* infection, renal cell carcinoma, and tight junction, were among those pathways enriched in the network-module.

We further processed the data using the Ingenuity Pathway Analysis (IPA), which was able to evaluate the results for both upregulated and downregulated phosphosites. First, we analyzed the data when the healthy individual samples were used as the control group. Here, we discovered a significant upregulation of protein phosphorylation in CKD (**Fig. 5D**), with the autophagy pathway being overrepresented. Individuals with CKD have altered body homeostasis, protein aggregation, inflammation, increased ROS production in mitochondria, and elevated oxidative stress (56). Therefore, the autophagy pathway is activated in response to inflammation. Many upregulated pathways in CKD were also upregulated in the high-grade clear cell RCC cohort. Interestingly, some of those upregulated pathways were related to cancer. Meanwhile, the apoptosis pathway, an established anti-cancer defense mechanism, was upregulated in high-grade clear cell RCC alone (57).

Second, we used CKD as the control group, observing similar patterns as before (**Fig. 5D)**. The organization of cytoskeleton and cytoplasm were upregulated in the high-grade clear cell but not in the low-grade clear cell RCC. Similar patterns were observed for microtubule dynamics and cancer of cells. When CKD was used as the control group, the difference between activated diseases and functions in the low-grade clear cell RCC samples became much more distinct due to the less pronounced immune response related to phosphosites, rationalizing our experimental design to have CKD patients as the control as well.

## DISCUSSION

Early diagnosis and monitoring of cancer through biofluids, such as blood or urine, has been a decades-long aim of medical diagnostics (58). Since protein phosphorylation is one of the most important and widespread molecular regulatory mechanisms that control almost all aspects of cellular functions, analysis of the phosphorylation network can conceivably provide clues regarding disease states (59). Traditionally, studying disease biomarkers from biofluids has been challenging due to the presence of phosphatases and high-abundant components in biofluids. However, the discovery of extracellular vesicles (EVs) opened new opportunities for developing phosphoproteins as potential cancer markers. Therefore, EVs have emerged as a rich resource for discovering tumor-relevant biomarkers from biofluids, such as urine, blood, etc. (60). Particularly promising are the discoveries that these EV-based disease markers can be identified well before the onset of symptoms or physiological detection of a tumor, making them promising candidates for early-stage cancer detection and disease diagnosis (61). In addition, EVs are membrane-covered nanoparticles, protecting the inside contents from external proteases and other enzymes (62–64). This attribute makes them highly stable in a biofluid for extended periods of time, a highly valuable feature for clinical settings. These features also make EVs a promising source for developing phosphoproteins as disease markers, considering many phosphorylation events directly reflect cellular physiological status. Together, these EV advantages allow EV phosphoproteins to be used as a promising source for RCC biomarker discovery.

In this project, we have developed a novel translational platform to isolate and profile phosphoproteins from circulating EVs for renal cell carcinoma grade differentiation. To achieve our overall goal, we explored the DIA method to differentiate between low-grade and high-grade clear cell RCC. DIA has become a powerful quantitative mass spectrometry technique, allowing researchers to identify and quantify time-sensitive phosphoproteomes in clinical samples (65). Here, we compared several DIA strategies to find the best suitable method for our mass spectrometry analysis to accomplish the objectives of our project. We found the 400-1100 m/z precursor range with forbidden zones, designed to lessen the quadrupole transmission edge effects, to be the most compatible method with our mass spectrometry system.

Furthermore, having a robust library is crucial for analyzing DIA data effectively. There are several ways to generate libraries, each with its own benefits and drawbacks (29). Moreover, phosphoproteome samples are highly dynamic. Considering all of the above, we decided to compare direct DIA and GPF-DIA as ways to generate the phosphoproteome library. These two were two efficient methods that are independent of available libraries for analyzing DIA data. When analyzing a single sample run, we showed that GPF-DIA gave higher identification than DDA analyzed by Sequest and the direct DIA. The GPF-DIA fractionated and simplified the samples during the LC-MS data acquisition at the quadrupole; therefore, GPF-DIA provided a more in-depth library than direct DIA and increased the identification number for a single sample run, as visualized in **Figure 1B**.

Interestingly, direct DIA and GPF-DIA for our main experiment both generated unique phosphosites. The simplified MS1 and MS2 data acquisitions in GPF-DIA were the main reason that this approach could generate unique phosphosites not found during the direct DIA analysis. On the other hand, some low-abundant phosphopeptides got diluted in the combined pool for GPF-DIA, giving rise to those unique phosphosites found only in direct DIA. We concluded that combining GPF-DIA and direct DIA phosphosites (4,057 phosphosites) would be valuable and would provide additional information about the cancer state. Indeed, we could find more relevant phosphosites that were overexpressed exclusively in low-grade or high-grade clear cell RCC EV samples. Some of the phosphosites visualized in **Figure 5A**, which could serve as grade-specific EV biomarkers, were only identified exclusively in GPF-DIA or direct DIA. Some of those phosphosites were already known to be involved in the clear cell RCC onset or progression. Our collective phosphosite data supported by prior literature offers a great opportunity for further validation studies to translate these potential non-invasive signaling markers.

The gene ontology analysis of the unique upregulated phosphosites showed that renal cell carcinoma pathways were upregulated only in the high-grade clear cell RCC group. These pathways included some already known phosphoproteins involved in renal cell carcinomas, such as PAK1, PAK2, PAK4, BRAF, and BAD (44, 50, 51). Along with the renal cell carcinoma pathway, ErbB signaling, proteoglycans, pathogenic *E. Coli* infection, renal cell carcinoma, and tight junction seen in **Figure 5B and 5C**, were among those pathways enriched clear cell RCC as shown in the tissue-based study (54). Meanwhile, elevated phosphorylation on SLC47A1, VPS37B, PDCD6IP, ARHGEF10L, PLEKHG3, and SOWAHC was only discovered in low-grade clear cell RCC (**Fig. 5A**). Based on the Protein Atlas data, these are known prognostic markers, favorable in renal cancer, and positively correlated with a higher survival rate. Therefore, it is expected to see the upregulation of those markers in the low-grade clear cell RCC compared to the high-grade staging. These low-grade markers could help urologists determine the best treatments and convince those newly diagnosed patients with low-grade clear cell RCC to undergo active surveillance instead of going straight to surgeries that could result in potential overtreatments and overspendings.

In conclusion, we have developed a strategy to utilize phosphoproteins in urinary extracellular vesicles for RCC grade differentiation. The study highlights our ability to isolate and identify thousands of unique phosphosites utilizing the EVtrap method and PolyMAC enrichment and combining the results of GPF-DIA and direct DIA analyses. These findings further promote the underlying principle that this novel strategy could be valuable for exploring existing resources in any diseases. Finally, we expect that our results, followed by extensive validation with a larger cohort of clinical samples, could significantly augment the practitioners assessment of a localized renal mass through a simple office based test and optimize management’ for many patients. Moreover, as a follow-up to this study, we have collected many additional samples with more diverse RCC subtypes and plan to apply the optimized method combined with an automation instrument for EV isolation to produce more reproducible results and validate those upregulated phosphosites uniquely found in low-grade or high-grade clear cell RCC. Overall, this project has created a translational environment where the collision of research with clinical perspectives has fostered the effective development of novel technologies and strategies to improve the effectiveness of cancer detection, diagnosis, and treatment.

## Data Availability

All data produced in the present study are available upon reasonable request to the authors.

## Acknowledgments

We thank the IU Health ECRO Biorepository for help in obtaining specimens used in this study. We also thank Thermo Fisher Scientific for providing our team with the Ultimate 3000 LC coupled to a Q-Exactive HF-X high-resolution MS instrument, which enabled this research. This project was supported by The Walther Embedding Program sponsored by the Walther Cancer Foundation, and NIH grant 3RF1AG064250. The cartoon in **Figures 1A and 2** was created with BioRender.com.

## Author Contributions

M.H., R.P., R.S.B., and W.A.T. conceived the project and designed the experiments. M.H. and Z.C.L. performed the main experiments. M.H., G.Z., Y.D., and H.Z. designed, compared, and optimized the DIA method. M.H. carried out the bioinformatics and statistical analyses and generated the figures, final data, and markers. M.H. and Z.L. designed and created **Figures 1A and 2** with BioRender.com. A.I. developed the protocols for sample normalization, the EVtrap, and PolyMAC enrichment experiments. M.H., Z.C.L., Z.L., A.I, and W.A.T. wrote the paper. All authors reviewed and approved the manuscript.

## Figure Captions

**Supplementary Table 1.** Second DIA method optimization. A random urine EV sample was used to compare between DIA without and with forbidden zones. The DIA data was searched in the class 1 library. The localization probability threshold of all phosphoproteome above is >0.00.

| DIA Method | All Class Phosphoproteome |
| --- | --- |
| DIA without forbidden zones | 670 / 1152 phosphoprotein groups (58% specificity) |
|  | 1956 / 4268 phosphopeptide groups (46% specificity) |
| DIA with forbidden zones | 685 / 1202 phosphoprotein groups (57% specificity) |
|  | 1987 / 4580 phosphopeptide groups (43% specificity) |

**Supplementary Table 2.** First DIA method optimization. A urine EV sample was used to compare DDA, direct DIA, and GPF DIA with 0.75 and 0.00 minimal localization thresholds. The direct DIA and the GPF DIA were performed at 3 different precursor ranges: 400-1000, 500-1100, and 400-1100 m/z.

| Search Method | Precursor Range (m/z) | Minimal Localization Threshold | The Number of Phosphoprotein | The Number of Phosphopeptides |
| --- | --- | --- | --- | --- |
| GPF | 400-1000 | 0.75 | 893 | 2484 |
| GPF | 400-1100 | 0.75 | 930 | 2660 |
| GPF | 500-1100 | 0.75 | 894 | 2489 |
| Direct DIA | 400-1000 | 0.75 | 588 | 1435 |
| Direct DIA | 400-1100 | 0.75 | 597 | 1449 |
| Direct DIA | 500-1100 | 0.75 | 571 | 1445 |
| DDA |  | 0.75 | 635 | 1176 |
| GPF | 400-1000 | 0 | 1179 | 4319 |
| GPF | 400-1100 | 0 | 1208 | 4680 |
| GPF | 500-1100 | 0 | 1186 | 4485 |
| Direct DIA | 400-1000 | 0 | 763 | 2435 |
| Direct DIA | 400-1100 | 0 | 778 | 2618 |
| Direct DIA | 500-1100 | 0 | 750 | 2441 |
| DDA |  | 0 | 721 | 1387 |

**Supplementary Figure 1.**
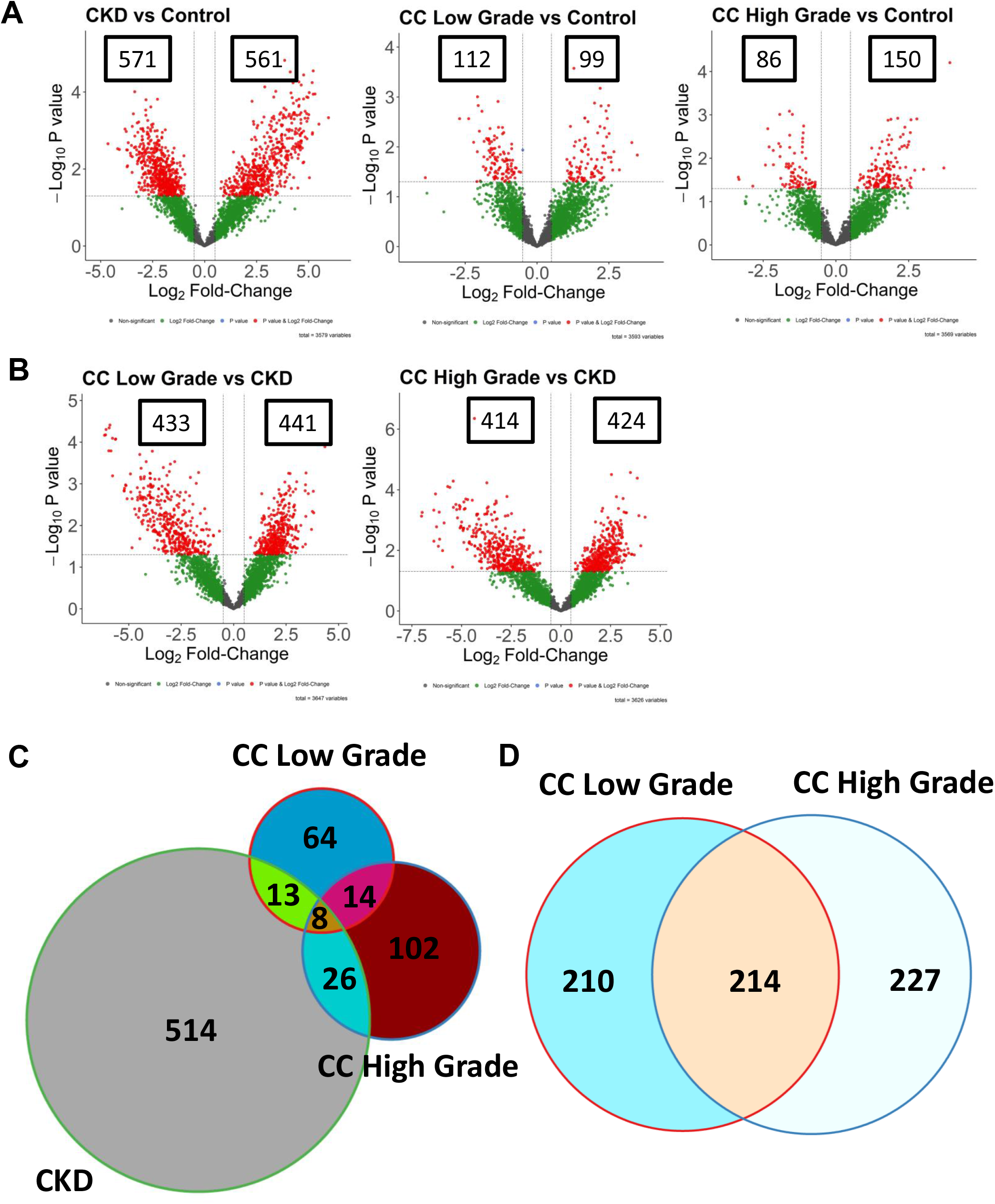
Upregulated phosphosite identification. A) All three groups: CKD, clear cell low-grade, and clear cell high-grade were compared to the Control group. B) Two groups: clear cell low-grade and clear cell high-grade were compared to the CKD group. Significantly up-regulated phosphosites compared to the control (C) and compared to the CKD (D) were overlapped in Venn diagrams. Volcano plots were created for each comparison with cut-off values of Welch’s two-sample t-test p-value = 0.05 and log base 2 difference = 0.5, which equals to ∼1.414 fold-change.

**Supplementary Figure 2.**
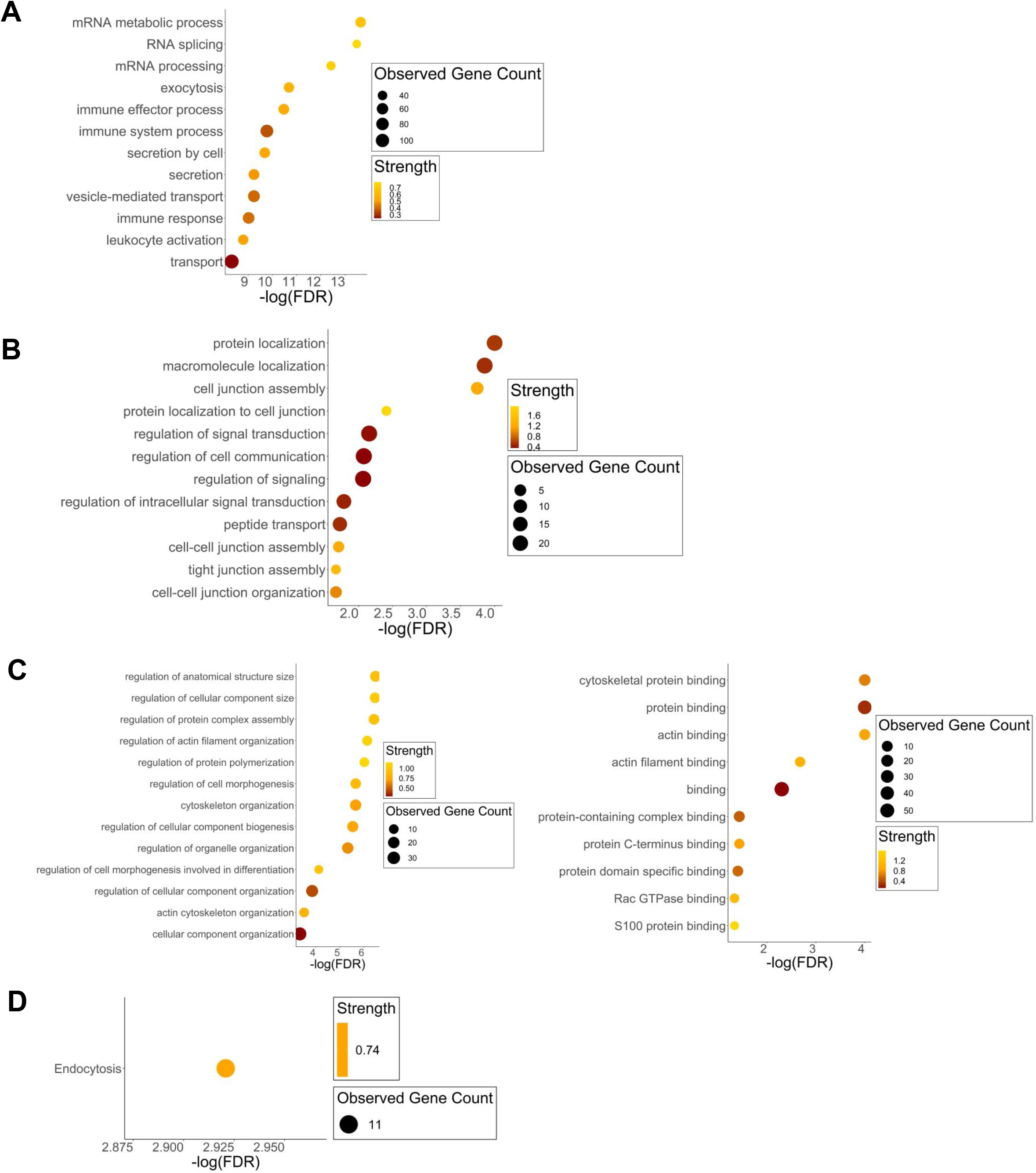
Enriched biological process gene ontology analyses of up-regulated phosphosites. The upregulated biological processes for A) CKD compared to control; B) clear cell low-grade compared to control; and C) clear cell high-grade compared to control (left) with the upregulated molecular functions for clear cell high-grade compared to control (right). D) The upregulated KEGG for clear cell low-grade compared to CKD. The analyses were carried out with the String database.

